# COVID-19 International Border Surveillance Cohort Study at Toronto’s Pearson Airport

**DOI:** 10.1101/2021.02.25.21252404

**Authors:** Vivek Goel, David Bulir, Eric De Propetis, Munaza Jamil, Laura Rosella, Dominik Mertz, Cheryl Regehr, Marek Smieja

## Abstract

**Objectives:** The primary objective was to estimate the positivity rate of air travelers coming to Toronto, Canada in September and October, 2020, at arrival, day 7 and day 14. Secondary objectives were to estimate degree of risk based on country of origin; to assess knowledge and attitudes towards COVID-19 control measures; and subjective well-being during the quarantine period.

**Design:** Prospective cohort of arriving international travelers.

**Setting:** Toronto Pearson Airport Terminal 1, Toronto, Canada.

**Participants:** Passengers arriving on international flights. Inclusion criteria were those aged 18 or older who had a final destination within 100 km of the airport; spoke English or French; and provided consent. Excluded were those taking a connecting flight; who had no internet access; who exhibited symptoms of COVID-19 on arrival; or who were exempted from quarantine.

**Main outcome measures:** Positive for SARS-CoV-2 virus on RT-PCR with self-administered nasal-oral swab, and general well-being using the WHO-5 index.

**Results:** Of 16,361 passengers enrolled, 248 (1·5%, 95% CI 1.3%,1.5%) tested positive. Of these, 167 (67%) were identified on arrival, 67 (27%) on day 7, and 14 (6%) on day 14. The positivity rate increased from 1% in September to 2% in October. Average well-being score declined from 19.8 (out of a maximum of 25) to 15.5 between arrival and day 7 (p<0.001).

**Conclusions:** A single arrival test will pick up two-thirds of individuals who will become positive, with most of the rest detected on the second test at day 7. These results support strategies identified through mathematical models that a reduced quarantine combined with testing can be as effective as a 14 day quarantine.

**Article Summary:** *Strengths and limitations of this study:*

- Decisions regarding border restrictions have been based on trial and error and mathematical models with limited empirical data to support such decision-making.
- This study assessed the prevalence of SARS-CoV-2 in a cohort of international travellers at arrival, day 7 and 14 of quarantine.
- It is limited to one airport and there is the potential from bias due to non-participation and loss to follow-up.
- Self-collected nasal-oral swabs were used which facilitated participation but may have reduced sensitivity.

## Introduction

As COVID-19 has rapidly spread across the globe and threatened the lives and safety of people in all regions, governments have attempted to find means to limit transmission, often relying on limited knowledge and evidence. Measures to control disease spread across international borders have included identification of ill passengers by symptom screening or temperature checks, strict quarantine requirements, or combinations of virus testing and quarantine. Many countries, such as Canada, have kept borders closed to foreign travelers, with the exception of essential workers and returning Canadians.^1^ This, coupled with a 14-day quarantine requirement, is designed to discourage international travel and minimize the risk of imported COVID-19 from abroad. Other countries have adopted strategies that either require a pre-departure test, arrival, and/or post arrival test combined with reduced or no quarantine. Some countries have also taken a risk-based approach, with varying requirements dependent on the risk of COVID-19 transmission in the origin country, or the activities the traveler will engage in. Decisions regarding such approaches have largely been based on trial and error or mathematical modelling.^2–7^ A recent Cochrane review concluded that the quality of evidence for most travel control measures was very low with mixed results, and that the optimal approach likely depends on a specific country’s context and own epidemiology.^8^

Quarantines can be difficult to enforce, have variable compliance, and may result in significant negative effects related to social isolation, restricted physical activity, lost productivity, and income.^9–11^ Direct impacts of border closures and travel hesitancy related to quarantine are felt in the travel industry which represents a large portion of the global economy.^12,13^

While testing of infected travelers should reduce the risk of disease importation, SARS-CoV-2 presents challenges given the potential for asymptomatic and pre-symptomatic transmission. Symptom screening and temperature checks will not detect asymptomatic or pre-symptomatic individuals. Testing, either pre-departure or on arrival will miss those individuals who have been just infected and are still incubating the virus. Thus, quarantine remains one of the few options, but the optimal length of quarantine remains unclear. One modelling study suggests that quarantine with testing on day 7 achieves a level of risk reduction similar to that of a 14-day quarantine.^14^ This assumes that there is perfect compliance and/or enforcement of the quarantine measures.

Given the substantial costs, and the impact of quarantine on personal well-being, it is critical to generate empirical data to support theoretical and mathematical models. To our knowledge, there are no systematic data reported on the proportion of international travelers that test positive for SARS-CoV-2 on arrival to Canada and during quarantine. The present study aimed to systematically estimate the positivity rate of air travelers coming to Toronto, Canada at arrival, day 7, and day 14 during a 2-month period in the fall of 2020. A further objective was to determine whether a combination of testing and country of origin could accurately identify those who were at highest risk for developing COVID-19. We also assessed knowledge and attitudes towards COVID-19 control measures and subjective well-being during the quarantine period.

## Methods

### Study Design

We conducted a prospective cohort study of arriving international travelers to Toronto, Canada Pearson International Airport, Terminal 1 between September 3, 2020 and October 31, 2020. The study protocol was reviewed and approved by the Advara Research Ethics Board.

Inclusion criteria were those aged 18 or older who had a final destination within 100 km of Toronto Pearson airport; could speak English or French; and provided consent. Exclusion criteria for the study were those passengers taking a connecting flight through Pearson Airport; who had no internet access; who exhibited symptoms of COVID-19 on arrival; or who were exempted from quarantine (e.g. essential workers).

### Enrollment Procedures

Individuals arriving on flights with participating Star Alliance airlines were invited to join the study during their flight or after arrival. Flights from any international destination that terminated at Pearson International Airport Terminal 1, Toronto, Ontario, were included. Flight crew announced the opportunity to participate in the study and directed passengers to view an instructional video that was prepared by the investigators in English and French on the in-flight entertainment systems. Flight crews were given a script to answer basic questions from potential participants, such as how to get to the study booth after landing, but referred interested individuals to investigator team’s research personnel for any questions related to the study. Information and study invitations were also posted in the arrivals and baggage areas of Terminal 1 in order to ensure that interested passengers could directly review and consider study materials after arrival.

Upon arrival, eligible and consenting passengers were guided to the testing booths. These were located in a secure area, after passage through immigration and customs, and following baggage collection. After reviewing a study information sheet and completing informed consent, participants proceeded to a specimen collection booth, where they were trained and supervised in the self-collection of oral-nasal swabs. Briefly, a flocked swab (Miraclean, Shenzhen, China) was moistened on the tongue, followed by bilateral swabbing of the buccal sulcus (between cheeks and gums) with rotation three times; followed by insertion into each nostril approximately 2-4 cm as a “deep nasal” swab and rotated three times. Swabs were placed in 2 ml of McMaster Molecular Medium, a guanidine isothiocyanate-based lysis buffer designed to inactivate virus and preserve RNA (Research St. Joseph’s, Hamilton Ontario). Participants were given 2 further specimen sampling kits for self-administration on day 7 and 14. For the two remaining tests after arrival, couriers were arranged to pick up the kits at the passengers’ location of quarantine.

The participants completed online questionnaires at the same three time points. Prompts were provided by SMS/text message to complete the follow-up tests and questionnaires. The questionnaires covered travel history, symptoms, mental health, attitudes towards protection measures (e.g., quarantine), and behaviours (e.g., handwashing). The items were drawn from the World Health Organization Survey Tool and Guidance for Rapid, Simple, Flexible Behavioural Insights on COVID-19.^15^

### Laboratory Methods

Swabs collected in McMaster Molecular Medium (MMM) were batch-processed from nucleic acid extraction to RT-qPCR set-up on the Hamilton Microlab® STAR (Hamilton Company, Nevada, USA). Nucleic acid extraction was performed using the Maxwell® HT Viral TNA Kit (Promega, Wisconsin, USA). For RT-qPCR, Luna® Universal Probe One-Step RT-qPCR (New England BioLabs, Massachusetts, USA) was used in combination with custom synthesized primers and probes (LGC Biosearch, California, USA). RT-qPCR was performed for 45 cycles on all specimens on the Bio-Rad CFX96 Touch Real-Time PCR Detection System (Bio-Rad Laboratories, California, USA) using the laboratory developed triplex assay, which contains two SARS-CoV-2 targets (Envelope, 5’-untranslated region {5’-UTR}), and a human housekeeping/sample adequacy target, RNase P. All PCR testing was conducted at The Research Institute of St. Joseph’s in Hamilton, ON. As testing was done in a research lab, in order to ensure appropriate confirmation and reporting to public health those who were determined “non-negative” were then referred to a provincial government COVID-19 assessment centre for a nasopharyngeal swab. For clarity, those who tested “non-negative” in the research laboratory will be referred to as “positives” throughout the manuscript.

Viral load, as measured by the average Cycle threshold (Ct) during polymerase chain reaction for envelope-gene and 5’UTR, was divided a priori into high viral load (Ct<25 cycles), moderate viral load (Ct 25-35), or low viral load (>35 cycles). All viral loads were obtained from duplicate PCR measurements, and where an analyte was not amplified, a value of 45 was imputed.

### Data management and analysis

All participant information and lab results were stored in a secure cloud-based information system.

The main end-points were rate of travellers testing positive for the first time on arrival, day 7, and day 14. Exact 95% confidence intervals for the rates were calculated using the Clopper-Pearson exact method. The denominator for calculating positivity by timepoint was the total number of participants who registered a test for the respective timepoint (i.e., arrival, day 7, and day 14). In order to account for individuals who did not complete follow-up tests, two approaches were taken. First, a sensitivity analysis was conducted by assuming different positivity rates based on best- and worst-case scenarios for those lost to follow-up. We conducted a second analysis where we used inverse probability weighting (IPW) to adjust for potential selection bias associated with dropouts. Specifically, we developed two IPW models using logistic regression to assess differences between those that dropped out on Day 7 and Day 14 versus those that did not and adjusted for positivity rates using the weights.^16^ Regarding our IPW approach, as weighted estimates no longer follow a binomial distribution, bootstrap 95% confidence intervals were used instead.^17^

Baseline descriptive statistics and measures of independence were computed for key demographic and travel related variables. Country of origin was classified based on the European Union risk framework.^18^ This classifies countries as low, medium and high risk based on incidence and test positivity rates. Countries with insufficient information or a testing rate of less than 300/100,000 population per week are classified as grey. Daily data posted by the European Centre for Disease Control was used to assess each country’s risk on the participant’s arrival date.

The general well-being items of feelings of cheerfulness, calm, interest in one’s life, feeling active, and well rested over the previous two weeks were combined into an average score based on the response categories.

As some participants did not complete all items on the questionnaires we used a multiple imputation (MI) approach to impute missing values. MI uses logistic and multinomial logistic regression to create multiple datasets of predicted values and takes the average across datasets as the final imputed value. Variables used in this approach were: gender, age, continent of origin, mental health, risk category, and handwashing (a behaviour variable). In the case of missing country of origin data, a grouped imputation approach was utilized. Given the large variance of responses, multiple imputation was not possible. Therefore, groups of twenty travellers that arrived at the study booth at the same time were made around missing values and the most frequent country of origin for these groups were imputed. This approach assumes that registrants usually arrive in groups as they are recruited on their respective flights.

Given the low prevalence expected on the final test, essentially zero, we used the method outlined in Frank et al for assessing our power.^19^ A sample size of 10,000 completed day 14 tests was selected to be sufficient to rule out the true proportion being 100% greater than that observed.

## Results

### Study Population

A total of 16,361 passengers registered for the study and completed at least one test. Study participants arrived from all continents (except Antarctica) and represented all risk categories and age groups. The highest proportion of participants were those arriving from the Americas (56%), countries classified as “red” according to the EU risk classifications (69%), and younger and middle age groups (73% between the ages of 18 and 49) (Table 1).

**Table 1:**
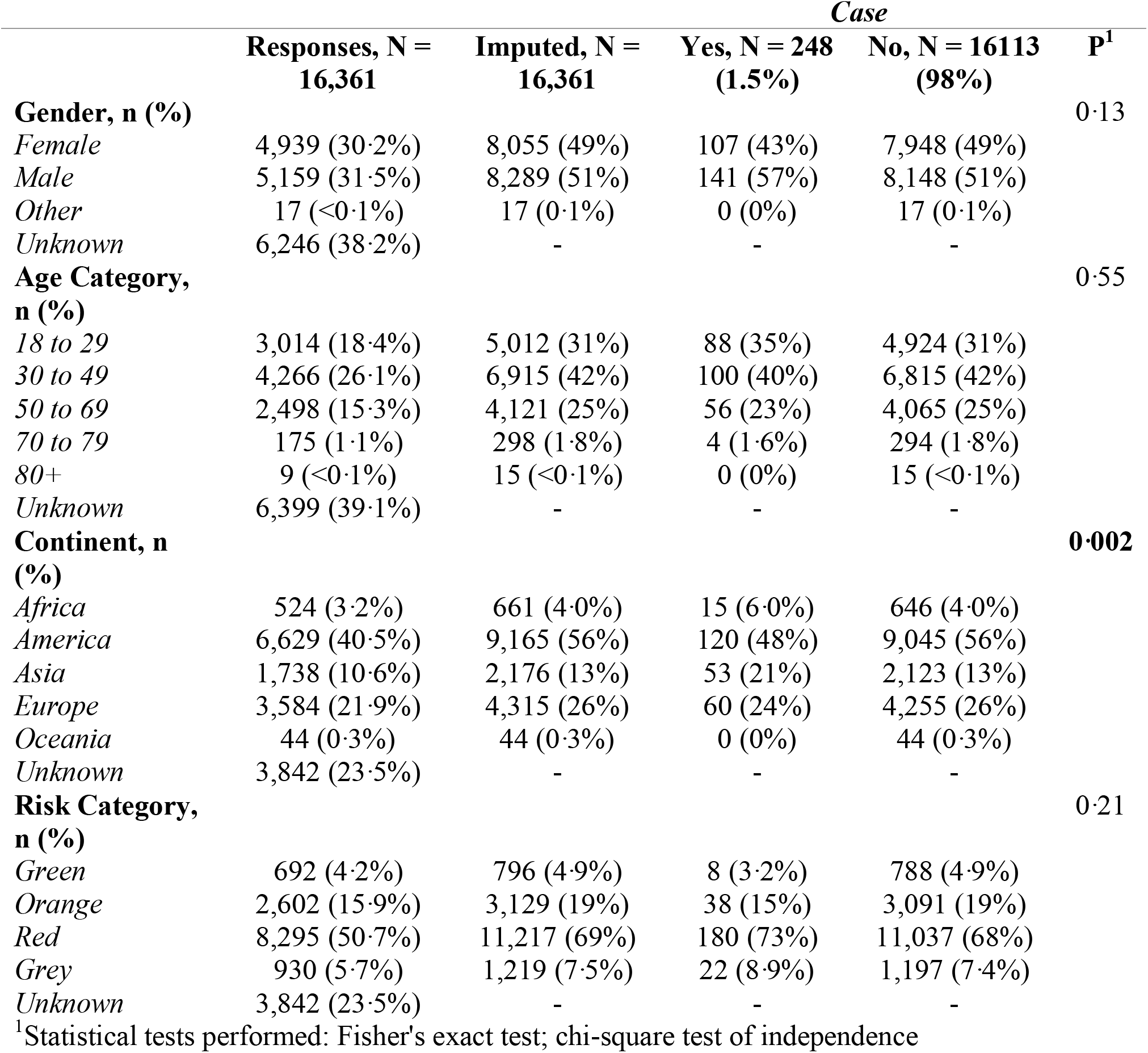
Baseline Characteristics by Case Counts.

Approximately 8 to 20% of the cohort was lost to follow-up at the various timepoints (Figure 1). Participants lost to follow-up were more likely to be in the youngest age group (36% between the ages of 18 to 29), arriving from the Americas (60%), or arriving from a red risk country (70%) (Table 2). The United States was over-represented as the country of origin among lost to follow-up participants.

**Figure 1:**
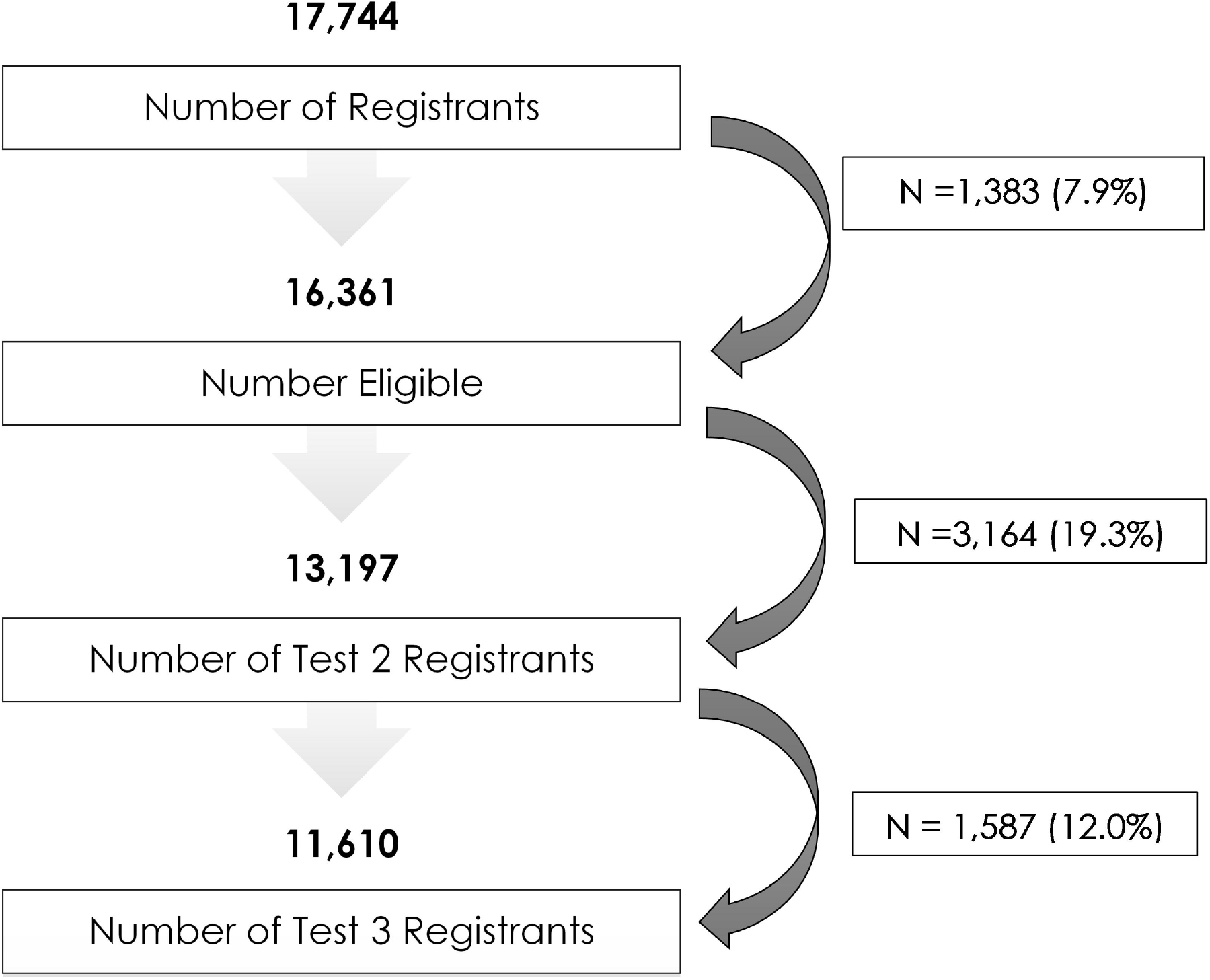
Participation Flow Chat.

**Table 2:** Demographics of Passengers Lost to Follow-up.

|  | **Responses, N =**<br>**16,361** | **Imputed, N =**<br>**16,361** | ***Loss to Follow-up*** |  |
| --- | --- | --- | --- | --- |
|  |  |  | **Loss, N = 4661**<br>**(28%)** | **No Loss, N =**<br>**11700<sup>1</sup> (72%)** |
| **Gender, n (%)** |  |  |  |  |
| *Female* | 4,939 (30.2%) | 8,055 (49%) | 2,237 (48%) | 5,818 (50%) |
| *Male* | 5,159 (31.5%) | 8,289 (51%) | 2,417 (52%) | 5,872 (50%) |
| *Other* | 17 (<0.1%) | 17 (0.1%) | 7 (0.2%) | 10 (<0.1%) |
| *Unknown* | 6,246 (38.2%) | - | - | - |
| **Age Category, n (%)** |  |  |  |  |
| *18 to 29* | 3,014 (18.4%) | 5,012 (31%) | 1,674 (36%) | 3,338 (29%) |
| *30 to 49* | 4,266 (26.1%) | 6,915 (42%) | 1,875 (40%) | 5,040 (43%) |
| *50 to 69* | 2,498 (15.3%) | 4,121 (25%) | 1,026 (22%) | 3,095 (26%) |
| *70 to 79* | 175 (1.1%) | 298 (1.8%) | 81 (1.7%) | 217 (1.9%) |
| *80+* | 9 (<0.1%) | 15 (<0.1%) | 5 (0.1%) | 10 (<0.1%) |
| *Unknown* | 6,399 (39.1%) | - | - | - |
| **Continent, n (%)** |  |  |  |  |
| *Africa* | 524 (3.2%) | 661 (4.0%) | 207 (4.4%) | 454 (3.9%) |
| *America* | 6,629 (40.5%) | 9,165 (56%) | 2,780 (60%) | 6,385 (55%) |
| *Asia* | 1,738 (10.6%) | 2,176 (13%) | 673 (14%) | 1,503 (13%) |
| *Europe* | 3,584 (21.9%) | 4,315 (26%) | 990 (21%) | 3,325 (28%) |
| *Oceania* | 44 (0.3%) | 44 (0.3%) | 11 (0.2%) | 33 (0.3%) |
| *Unknown* | 3,842 (23.5%) | - | - | - |
| **Risk Category, n (%)** |  |  |  |  |
| *Green* | 692 (4.2%) | 796 (4.9%) | 213 (4.6%) | 583 (5.0%) |
| *Orange* | 2,602 (15.9%) | 3,129 (19%) | 787 (17%) | 2,342 (20%) |
| *Red* | 8,295 (50.7%) | 11,217 (69%) | 3,285 (70%) | 7,932 (68%) |
| *Grey* | 930 (5.7%) | 1,219 (7.5%) | 376 (8.1%) | 843 (7.2%) |
| *Unknown* | 3,842 (23.5%) | - | - | - |

<sup>1</sup> Passengers who tested positive for COVID were counted as completing the study irrespective of how many follow-up tests they completed considering disease status is known.

### Symptoms and Positive COVID-19 Results

Throughout the study period, 248 (1·5% of those completing the first test, 95% CI 1.3%,1.5%) individuals who tested positive at least once were identified. Of these cases, 167 (67%) were identified on arrival, 67 (27%) on day 7, and 14 (6%) on day 14 of quarantine (Table 3). The proportion of positive participants increased from early September to end of October (Figure 2).

**Table 3:** Primary Results.

| **Time** | **Cases** | **N** | **Rates**<br>**per**<br>**100k** | **Lower**<br>**CI** | **Upper**<br>**CI** |
| --- | --- | --- | --- | --- | --- |
| Overall | 248 | 16,361 | 1,516 | 1,334 | 1,715 |
| Arrival | 167 | 16,361 | 1,021 | 872 | 1,187 |
| Day 7 | 67 | 13,197 | 508 | 394 | 644 |
| Day 14 | 14 | 11,610 | 121 | 66 | 202 |

**Figure 2:**
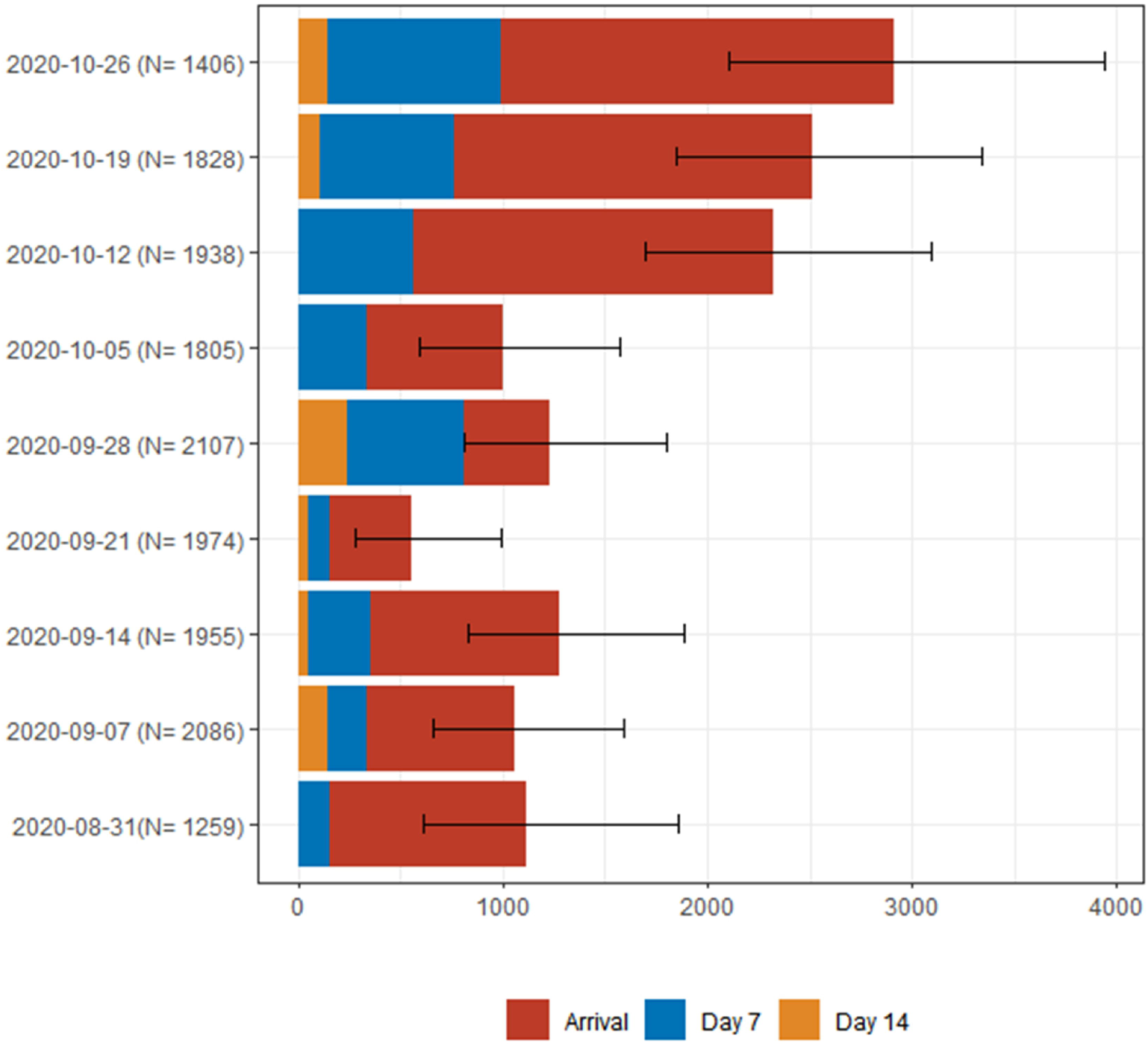
Rates of COVID-19 Infections by Week of Arrival. Note: Test numbers indicate rates for when passengers first became positive for COVID-19.

Of the 167 individuals who tested COVID-19 positive on arrival, 3 participants reported symptoms at arrival, 30 participants at day 7, and 14 participants at day 14 (Table 4). At each time period, a higher proportion of those who tested positive were asymptomatic or pre- symptomatic.

**Table 4:**
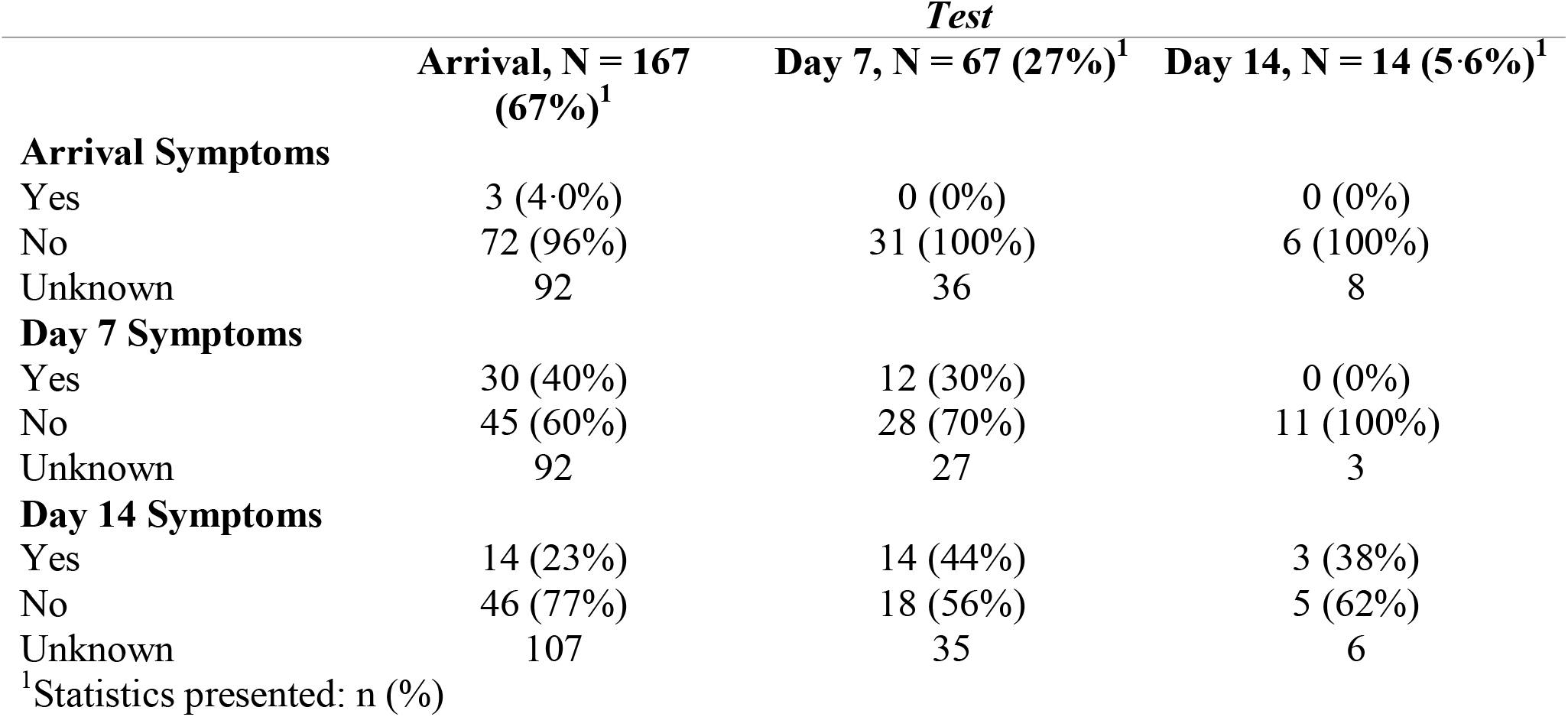
Symptomatic Status of Positives by Timepoint.

Passengers who tested positive during the study period were more likely to be in the younger age groups, male (57%), and to be arriving from countries classified as “red” in terms of risk (Figure 3). In comparison to those who tested negative during the study period, the highest proportion of positive travelers came from “grey” risk countries (7·4% vs 8·9%).

**Figure 3:**
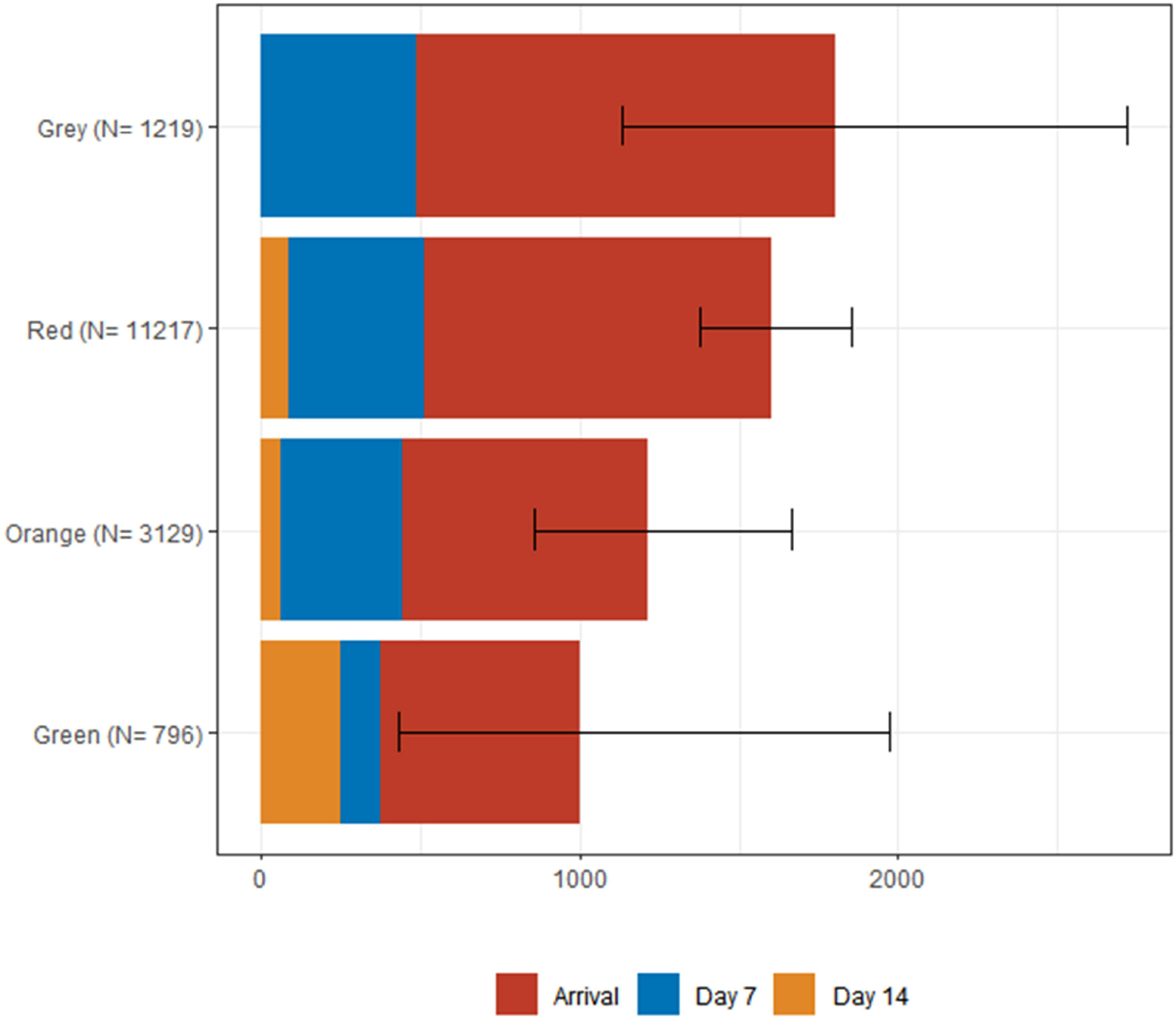
Rates of COVID-19 Infections by ECDC Risk Category. Note: Test numbers indicate rates for when passengers first became positive for COVID-19.

In order to address the possibility of missed cases from the travellers who were lost to follow-up, 1% of these losses were added to the overall case counts for day 7 and day 14. After this analysis was run, the conservative case count estimate for the entire study period was 292 with 107 of these cases being picked up on day 7 and 18 on day 14 (Table 5). Similar results were obtained with the inverse probability weights method.

**Table 5:** Sensitivity Analysis to Address Loss to Follow-up.

**Adding One Percent of Those Lost to Follow-up**
| **Time** | **Cases** | **Rate/**<br>**100k** | **Lower**<br>**CI** | **Upper**<br>**CI** |
| --- | --- | --- | --- | --- |
| Overall | 292 | 1,785 | 1,587 | 1,999 |
| Arrival | 167 | 1,021 | 872 | 1,187 |
| Day 7 | 107 | 654 | 536 | 790 |
| Day 14 | 18 | 110 | 65 | 174 |

**Inverse Probability Weighted Estimation**
| **Time** | **Cases** | **Rate/**<br>**100k** | **Lower**<br>**CI** | **Upper**<br>**CI** |
| --- | --- | --- | --- | --- |
| Overall | 272 | 1662 | 1455 | 1858 |
| Arrival | 167 | 1021 | 862 | 1179 |
| Day 7 | 84 | 519 | 404 | 627 |
| Day 14 | 21 | 130 | 73 | 185 |

### Viral Load Estimation

The level of viral load in positive cases is presented in Table 6. Low viral load was present on arrival in 60 out of 137 (36%) positive individuals, 20 out of 67 (30%) at day 7, and 6 out of 14 (43%) at day 14. Only 2 of the 14 positives on day 14 had a high viral load.

**Table 6:** Cycle threshold (Ct) as surrogate for RNA load by First Positive Test.

|  | *Test* |  |  |
| --- | --- | --- | --- |
|  | Arrival, N = 167<sup>1</sup> | Day 7, N = 67<sup>1</sup> | Day 14, N = 14<sup>1</sup> |
| **Average Ct\* Across All Targets** | 32 (26, 38) | 27 (22, 36) | 30 (27, 37) |
| **Viral Load** |  |  |  |
| Low (Ct>35) | 60 (36%) | 20 (30%) | 6 (43%) |
| Moderate (Ct 25-35) | 71 (43%) | 21 (31%) | 6 (43%) |
| High (Ct <25) | 36 (22%) | 26 (39%) | 2 (14%) |

<sup>1</sup>Statistics presented: Median (IQR); n (%)

Well-being and Attitudes Regarding COVID-19 and Public Health Approaches

Figure 4 presents the change in mental health score by time point. Participants had a much more favourable disposition on arrival than during quarantine.

**Figure 4:**
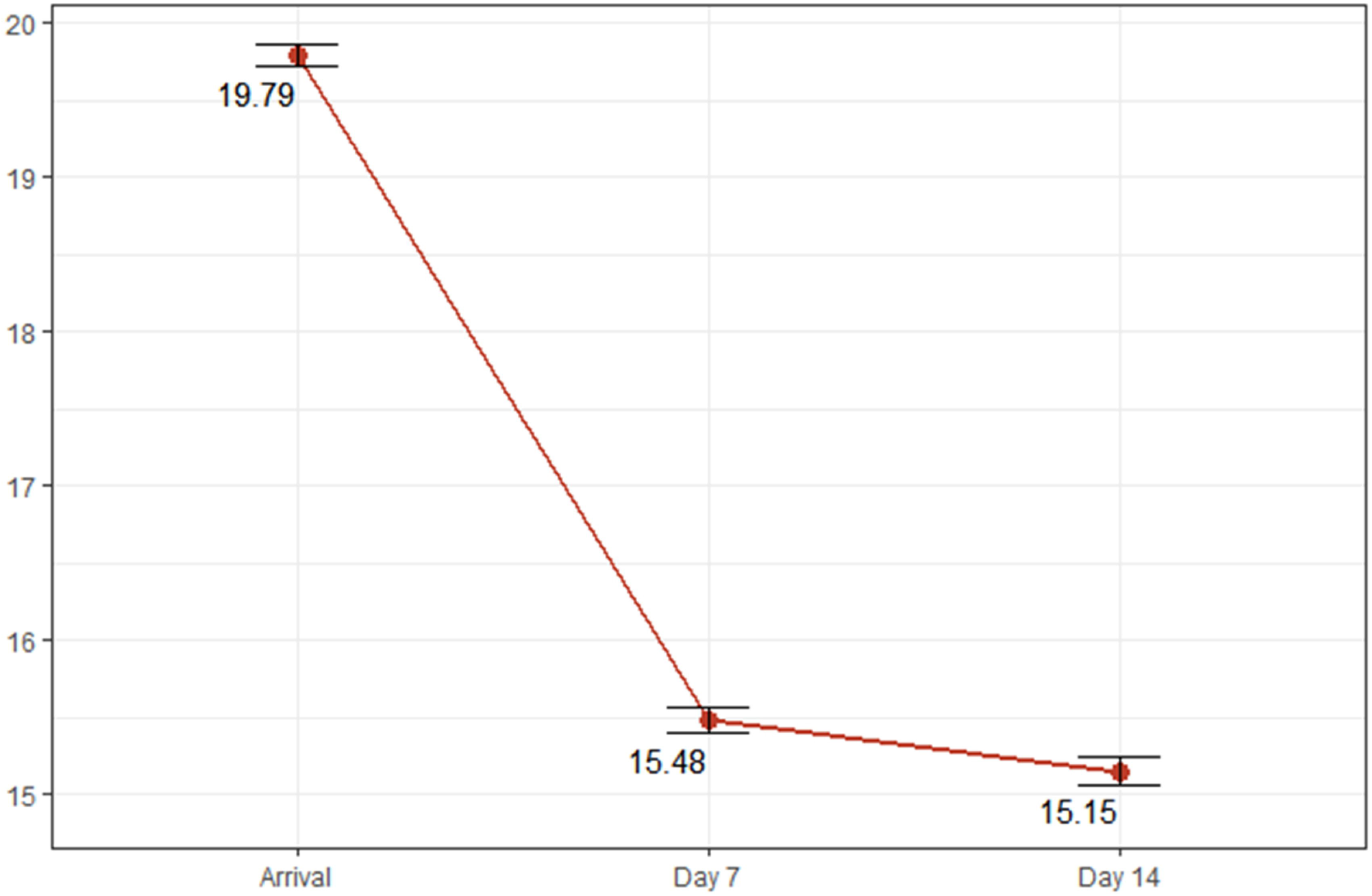
Average Mental Health Score by Day of Quarantine. The maximum possible score is 25. P<0.001 for decline in score from arrival to Day 7 and 14.

Table 7 presents two key attitudinal items regarding controls for international travel. Participants were more willing to accept a requirement for testing than for immunization.

**Table 7:** Baseline Attitude Responses.

|  | *Case* |  |
| --- | --- | --- |
|  | **Yes, N = 248<sup>1</sup>** | **No, N = 16,113<sup>1</sup>** |
| **COVID Test Required<sup>2</sup>** |  |  |
| very acceptable | 54 (50%) | 5,513 (57%) |
| acceptable | 40 (37%) | 3,021 (31%) |
| neither acceptable nor unacceptable | 7 (6.5%) | 804 (8.2%) |
| unacceptable | 5 (4.7%) | 285 (2.9%) |
| very unacceptable | 1 (0.9%) | 132 (1.4%) |
| Unknown | 141 | 6,358 |
| **Vaccination Required<sup>3</sup>** |  |  |
| very acceptable | 33 (31%) | 3,685 (38%) |
| acceptable | 33 (31%) | 2,631 (27%) |
| neither acceptable nor unacceptable | 25 (24%) | 1,615 (17%) |
| unacceptable | 7 (6.6%) | 928 (9.6%) |
| very unacceptable | 8 (7.5%) | 828 (8.5%) |
| Unknown | 142 | 6,426 |

<sup>1</sup>Statistics presented: n (%)
<sup>2</sup> “If a negative COVID-19 test were required for international travel in the future how acceptable would you find that”?
<sup>3</sup> “If proof of a COVID-19 vaccination were required for international travel in the future, how acceptable would you find that”?

## Discussion

In order to control the spread of COVID-19 globally, governments across the world are resorting to the use of travel restrictions and quarantine. In today’s globalized environment in which travel is ubiquitous and central to many national economies, it is critical to determine which measures are most appropriate. This study aimed to systematically estimate the positivity rate, symptoms, attitudes, and well-being of travelers on arrival in Toronto, Canada and during their 14-day quarantine in the fall of 2020.

Results revealed that 1·5% of study participants arriving as international passengers at Toronto Pearson airport tested positive by RT-PCR on or after arrival. The overall rates were approximately 1% in September and 2% in October, reflecting the rapidly changing conditions in the United States and Europe, which were the regions of origin for the largest proportion of arriving passengers. Approximately two-thirds of positive cases were detected on arrival, with most of the remaining cases being identified on day 7. Of the small number that were positive on day 14, approximately half had very low viral loads suggesting that their positive status may have been an artifact of previous infection and did not reflect active infection.

Males had higher positivity rates, although not statistically significant, than females. Preliminary analysis suggests that males reported being less likely to comply with public health recommendations (data not presented).

These results support those from modelling studies and the recent recommendations from the United States Centers for Disease Control that a shortened quarantine period of 7 days combined with a negative test would provide the same degree of control as a 14 day quarantine.^20^ This is particularly important given the significant impact observed on general well-being among the participants – consistent with other studies.^9^ However, such a recommendation assumes perfect compliance with quarantine which is difficult to achieve unless very strict requirements, such as specialized quarantine facilities, or GPS tracking, are implemented.^11^ It is plausible that compliance with a 7 day quarantine and test regimen may be better than a 14 day quarantine.

A relationship with risk of COVID-19 in the origin country was observed. However, the differences between risk categories was small and the benefits of a risk-based approach may be marginal. Furthermore, countries move rapidly through the different risk categories which makes implementation of such approaches difficult.

Russell et al^21^ have modelled when travel restrictions might have greatest impact. They show that stringent restrictions will be most effective for countries with low COVID-19 incidence and large numbers of arrivals from other countries, or where epidemics are close to tipping points. They recommend that countries should consider local COVID-19 incidence, local epidemic growth, and travel in determining appropriate restrictions.

In assessing whether or not to use testing to replace or reduce quarantine another important consideration is the availability of testing. In settings where availability of testing is limited, prioritizing border testing in order to reduce quarantine time may not be warranted. However, with the emergence of variants of concern, surveillance testing at borders may need to be increased. Indeed, many countries have tightened border controls including quarantine and testing requirements following the identification of variants of concern.

### Limitations

This study was conducted at a single terminal at Toronto’s Pearson Airport, albeit representing the majority of international flights arriving at Canada’s busiest airport. We enrolled approximately 20% of passengers. We believe that up to one-half of the arriving passengers would have met our exclusion criteria, so our participation rate likely approached 40%. There will likely have been selection bias in our participants. However, it is uncertain what the direction may have been. It may be possible that those who engaged in higher risk behaviours while abroad may have chosen not to participate. On the other hand, during the study period, PCR testing was not broadly available in Ontario, and some participants told us they took part in order to access the free testing. Regardless, selection bias would affect the overall positivity rate. However, a key value of our results is the distribution of positivity across the three timepoints, which should not be affected by selection bias.

We had losses to follow-up, and these may have biased the results if those who broke quarantine or developed symptoms might have been less likely to participate, or less likely to follow-up. We adjusted using a sensitivity analysis and inverse probability weighting, with similar results using both methods. IPW were based only on the variables that were measured and thus selection bias due to unmeasured factors may not be accounted for. The overall conclusions are not changed with either method.

We used supervised self-collection at the airport, and unsupervised self-collection at home for follow-up. Measurement of RNaseP levels found inadequate levels in only 0.2%, suggesting the approach enabled adequate sample collection in the vast majority of participants. The correlation between oral-nasal self-collection and staff-administered nasopharyngeal collection is estimated at 90-95% sensitivity, and the use of serial weekly collection on three occasions over 14 days of quarantine may have increased sensitivity.

## Conclusions

We demonstrated the feasibility of large-scale self-collection of oral-nasal swabs, coupled with a highly-sensitive, laboratory-based PCR testing, for arrival and follow-up testing of international passengers. Approximately 94% of infections were identified through arrival and day 7 testing, confirming findings from mathematical models that a 7-day quarantine coupled with testing would be highly effective in identifying importation of COVID-19. Given concern about importation of spike-protein variants of potential public health significance, airport testing would enable more timely detection and tracing of imported COVID-19 variants.

## Supporting information

STROBE Checklist

## Data Availability

Deidentified participant data will be made available with publication through an open data repository.

## Author Contributions

All authors contributed to the development and implementation of the study, review of results and preparation of the manuscript. MS and DB supervised the laboratory analysis. VG, LR and ED conducted the data analysis and verified the underlying data.

## Funding Acknowledgements

The conduct of the study was protected from the role of the funders. McMaster Health Labs (MHL) is a not-for-profit corporation created by McMaster University and Research St. Joseph’s to conduct COVID-19 related lab research. The contracts for funding with Air Canada and the Greater Toronto Airport Authority were with MHL. The funders also provided in-kind support for recruitment of participants and the study site at the airport. The Canadian Institutes for Health Research awarded a peer-reviewed grant for conduct of the study. The funders did not have any direction in the study design, execution, data analysis, or preparation of the manuscripts.

